# Trajectories of Cognitive Function in First-Episode Psychosis: Associations to Clinical Outcomes and Biomarkers

**DOI:** 10.1101/2024.12.03.24318204

**Authors:** Maria Lee, Alexis E. Cullen, Granville J. Matheson, Zheng-An Lu, Sarah E. Bergen, Carl M. Sellgren, Sophie Erhardt, Helena Fatouros-Bergman, Simon Cervenka

## Abstract

**Aims:** Cognitive dysfunction in psychotic disorders is common. At disorder onset, this impairment varies greatly between individuals, which may reflect different levels of decline compared to pre-morbid levels. Diverse trajectories in cognitive change prior to psychosis onset have been hypothesized to represent different underlying pathological processes. Our primary aim was to model cognitive change over time in a sample of individuals with first-episode psychosis (FEP) and controls. The secondary aim was to explore whether cognitive change was associated with clinical outcomes, and biological markers that have shown associations with disease progression.

**Methods:** Our sample consisted of 73 individuals with FEP who had undergone cognitive assessment at psychosis onset and 53 controls. Using school grades from registry data as a proxy for pre-morbid cognitive ability, we modelled change in cognition using linear mixed-effects models. The resulting change scores were correlated to polygenic risk scores, cerebrospinal fluid levels of complement protein C4A and clinical outcomes.

**Results and Conclusions:** Groups did not differ in school performance prior to psychosis. Psychosis onset was associated with a large cognitive decline in FEP and thereafter they performed significantly worse than controls. Among FEP individuals, there was a large degree of variability in cognitive change leading up to psychosis onset. Degree of cognitive change was not associated to the selected biological variables but did predict worse clinical outcomes. The results indicate that individual cognitive trajectories may be a clinically relevant topic for further study, but given the exploratory nature of the analysis, replication in an independent sample is required.

## Introduction

Cognitive impairment is an important symptom dimension of psychotic disorders such as schizophrenia ^1–3^, and has been related to functional impairment both at illness onset ^4,5^ and in later phases of the disorder ^4,6,7^. Whilst cognitive impairment was initially suggested to reflect an underlying neurodegenerative process ^8^, the neurodevelopmental model of psychotic disorders has since prevailed, supported by studies showing that cognitive problems emerge before the onset of positive and negative psychotic symptoms, with mild cognitive deficits evident already in childhood ^9–11^. How cognition develops in the years leading up to psychosis, and whether this differs between individuals, are questions of great importance, as this knowledge may provide insights into underlying pathological processes.

The most common approach to studying cognitive change is to estimate pre-morbid cognitive ability using reading tests such as the National Adult Reading Test (NART) ^12^, where single irregular words are read aloud. Reading tests have good psychometric properties ^13^, and can provide an estimate of cognitive abilities prior to illness onset. Investigations using reading tests combined with demographic factors find that a large proportion of patients with psychosis perform at a level significantly below their predicted premorbid abilities ^14,15^. However, these tests are not available in all contexts and ultimately, only provide estimates and not actual longitudinal measurements.

Establishing more precise trajectories of cognitive function in the years prior to psychosis onset requires prospective cohort studies or follow-back studies, both of which are rare. In one such prospective cohort study, using longitudinal cognitive data from infancy (18 months) until 20 years of age, individuals with psychotic disorders (n = 16) exhibited a mean decline in full-scale IQ of 1.09 standard deviations (SD) ^16^. Two Nordic longitudinal cohort studies in males demonstrated a cognitive decline relative to peers between the ages of 12-13 and 18 ^17,18^, in those who later went on to develop schizophrenia. In both studies, the degree of cognitive decline was an independent predictor of developing schizophrenia.

In clinical practice, pre-morbid cognitive ability is more commonly assessed using school grades and academic achievements. Although school grades are not identical to results from cognitive tests, cognitive ability has consistently been found to be the strongest predictor of scholastic achievement ^19,20^ with correlation coefficients of up to 0.7 reported ^19^. Furthermore, school grades are readily accessible to both clinicians and researchers. Consistent with reports that lower pre-morbid cognitive function is already present at the point of illness onset, individuals who go on to develop psychotic disorders have been found to exhibit lower scores on scholastic achievements tests compared individuals that do not ^21^, and lower scores compared to state norms ^22^. In these studies, patients were found to display a drop in performance in the ages 13-16 ^22^, as well as a widening gap compared to controls over time ^21^. Moreover, in a recent meta-analysis, general academic achievement before age 16 was found to be significantly lower in those who went on to develop schizophrenia compared to those who did not, although effect sizes were small (Cohen’s *d* =-0.29) ^23^.

In line with the general shift towards precision medicine approaches, there is an increased focus on investigating interindividual variability in cognitive function. Cognitive performance in psychotic disorders differs greatly between individuals ^24^, and this inter-individual variability was recently demonstrated to be significantly greater in patients than in healthy controls ^25^. Whether variability at illness onset reflects differences in trajectories rather than different pre-morbid levels of cognition is poorly understood. If trajectories do differ, what are potential biological, psychosocial, demographic or clinical factors associated with different magnitudes of change? Factors related to the longitudinal course of cognition could potentially be different from those associated with cognitive performance at psychosis onset.

Prior studies on factors related to cognitive trajectories have mainly focused on the discrepancy between reading tests of pre-morbid ability and current cognitive function, for instance linking early cognitive impairment to higher polygenic risk scores (PRS) for schizophrenia ^26^, or showing that the degree of deviation from expected pre-morbid ability is related to functional outcome and neurophysiological measures ^14^. Whilst these studies suggest that cognitive trajectories are associated with treatment response and other outcomes, it is important to acknowledge that they did not utilize longitudinal measurements.

Cohort studies with longitudinal data have used the degree of cognitive change to predict risk of developing schizophrenia ^27–30^, but to our knowledge, inter-individual variability in cognitive change has not been quantified previously.

In the present study we sought to extend prior research by using longitudinal data acquired before individuals developed their first psychotic episode. Using a well-characterized cohort of FEP individuals, our aims were to examine the between-individual variability in the magnitude of cognitive change prior to psychosis onset, and to relate the degree of cognitive change to subsequent health care usage (days in hospital and number of hospital admissions). These variables have previously been shown to be linked to cognitive function ^31^ but, to our knowledge, have not been examined in relation to cognitive change before psychosis onset. A further aim was to explore potential relationships between the degree of cognitive change and a candidate biological marker for psychosis in the form of the immune signaling protein complement component 4 A (C4A) as well as PRS for schizophrenia and intelligence.

## Methods

### Participants

Individuals with FEP were recruited through the ongoing Karolinska Schizophrenia Project (KaSP), which enrolls patients who experience their first psychotic episode and are either naïve to antipsychotic medication or within four weeks of first exposure to such medications. Inclusion criteria for patients was a diagnosis of psychotic disorder (except for substance-induced psychotic disorders and those related to a general medical condition) according to Structured Clinical Interview for DSM-IV (SCID)^32^, conducted by a psychiatrist or clinical psychologist. Controls were recruited through advertisement. Exclusion criteria for all participants were: severe somatic illness or neurological disorder and current abuse of alcohol or illegal drugs (including cannabis) or a history of such abuse. Further exclusion criteria for controls were current psychiatric illness assessed by Mini International Neuropsychiatric Interview (MINI) ^33^, lifetime use of antipsychotics or any first-degree relative with psychotic illness.

For the present study, participants were included if they had data on at least one cognitive measurement and registry data on school grades from at least one time-point. The final analytic sample consisted of 72 individuals with FEP and 53 controls.

### Data collection

As part of KaSP, individuals with FEP undergo clinical characterization (with the Positive and Negative Syndrome Scale (SCI-PANSS)^34^, Global Assessment of Function (GAF)^35^, Clinical Global Impression Scale (CGI)^36^). All participants in the study underwent cognitive testing with the MATRICS Consensus Cognitive Battery (MCCB) ^37^, lumbar puncture and blood sampling. The MCCB is the gold standard cognitive battery in schizophrenia research and shows high correlations with other measures of general intelligence, such as the Wechsler Abbreviated Scale of Intelligence Estimated Full Scale IQ ^38^. The MCCB consists of 10 subtests, measuring seven domains: Processing Speed, Working Memory, Verbal Learning, Visual Learning, Reasoning/Problem-solving, Attention and Social Cognition. For this study, we chose the global neurocognitive composite as our outcome measure, which was deemed to correspond better to general academic achievement. This measure includes nine out of the 10 subtests, excluding the social cognitive domain. Cognitive test scores were derived using the MCCB norms, with age and sex corrections applied. American norms were used, shown to be feasible in Scandinavian populations^39^.

### School grades

The Swedish National Agency for Education provides individual-level data on school grades^40^. Data were available from year 9 (ages 15-16 years), which marks the end of compulsory school in Sweden and from year 12 (ages 18-19 years), which is the end of upper secondary school. Approximately 75-80% of students who complete year 9 also complete year 12 (based on population level statistics).

### Health care use

The National Patient Register (NPR) ^41^records all inpatient visits to hospital, as well as outpatient visits to medical doctors in specialised care. The register includes data on diagnostic codes, duration of hospital stay, type of psychiatric care (voluntary, involuntary and mandated involuntary care in outpatient settings, referred to in Swedish as “öppen psykiatrisk tvångsvård”), and healthcare interventions. Registry data on health care use for study participants was collected from time of inclusion in study until 2022-09-30.

### Medication use

The Swedish National Prescribed Drug Register (PDR) ^42^ records all dispensations of prescribed medications from pharmacies throughout the country (not including medications administered in inpatient settings). The register includes data on date of prescription and dispensation, active substance, packaging and dosage. Registry data on medication dispensation for study participants was collected from time of inclusion in study until 2022-09-30.

Clinical outcomes were defined using a range of registry variables. For inpatient health care use, we calculated the number of hospital days (excluding mandated involuntary care in outpatient settings) and the number of hospitalizations due to any psychiatric diagnosis or self-harm (see Supplementary Material for exact definition) over the entire follow-up period (inclusion until 2022-09-30). To capture the need for continued psychiatric care and anti-psychotic medication following the first psychotic episode, we chose to focus on long-term clinical variables: outpatient psychiatric care in the fifth year after inclusion in study and dispensation of antipsychotic medication during the same time period. This was determined by outpatient visit in psychiatric care (12-month period prevalence prior to the 5-year mark of inclusion in study) with a main diagnosis for (1) any psychiatric diagnosis, (defined as International Classification of Diseases ^43^ (ICD) codes F10-F19, F30-F99), and (2) a main diagnosis corresponding to schizophrenia spectrum and other psychotic disorders (ICD codes F20 to F29). The same principle was applied for dispensation of antipsychotic medications (not including mood-stabilizers, for a full list see Supplementary Material).

### Polygenic Risk Scores

PLINK version 1.90 was employed to perform quality control of genetic data and generate PRSs ^44^. The base datasets for schizophrenia and intelligence PRSs are the summary statistics for the largest published GWAS ^45,46^ for these phenotypes. The target dataset for PRS calculation is the genotype data (provided using the InfiniumPsychArray-24 v1.1 BeadChip) from the study sample consisting of 93 individuals. The SNPs were excluded if they: (1) showed deviations from Hardy-Weinberg equilibrium (P < 1 × 10^-6^) or; (2) had a genotype missing rate > 5%; or (3) had a minor allele frequency < 1%, leaving a total of 318, 915 SNPs. Individuals with a genotype missing rate > 5% (N = 3) were excluded.

Quality control for the base datasets was performed by removing multiallelic and duplicated SNPs. After that, the target and base datasets were harmonized by extracting SNPs that were included in both datasets and aligning the effect alleles. After clumping the SNPs at r^2^ < 0.1 within 250 kb, a total of 58,829 and 54,281 SNPs were included as valid predictors for the schizophrenia and intelligence PRSs, respectively.

The PRSs were generated using the PRS-principal component analysis (PCA) approach^47^. First, the PRS at 10 different p-value thresholds (5e-8, 1e-6, 1e-4, 0.001,0.01, 0.05, 0.1, 0.2, 0.5, 1) were calculated. Next, PCA was applied to PRSs at all thresholds. The standardized first components derived from PCA were utilized as the final PRSs tested for associations with cognitive change. This consolidates risk from the individual PRSs, resulting in a single measure that is more powerful than a PRS at any specific p-value threshold and eliminating the need for multiple tests across thresholds.

### C4A protein

C4A protein levels were measured in cerebrospinal fluid (CSF) using targeted mass spectrometry and the Micro BCA Protein Assay Kit. Measurements were performed in a 96-well plate and fluorescence read by plate reader FLUOstar Omega. Sample preparation was performed on the Agilent AssayMAP Bravo Platform. Further details regarding sample collection and data analysis have been published previously ^48^.

### Statistical analysis

Individual participant grades were converted to percentiles by comparing their results to percentile values for the entire population that graduated in Sweden that year (data provided by the Swedish National Agency for Education). This was performed for grades from year 9 and year 12, respectively. The percentile scores were then transformed to their corresponding z-scores: a one-to-one independent transformation which maps the scores from a bounded uniform to a normal distribution, more accurately summarizing the distances between extreme values, and allows the use of parametric statistical models (for details of this procedure and flow chart see Supplementary Material).

For the MCCB, one FEP individual was missing results for one (Continuous Performance Test – Identical Pairs) of the 9 subtests used to derive the neurocognitive composite score. Imputation of this value was performed via stochastic regression using the mice package ^49^, in R, following the recommended methods described in the MCCB manual ^50^. The MCCB software produces percentile values, which were converted to z-scores in the same 1 to 1 sample independent transformation as for grades.

Linear mixed-effects modelling (LME) was used to explore cognitive trajectories using data from all 4 time-points. Models were built in consecutive steps, starting with random intercept for all participants, thereafter adding in turn: 1) timepoint, 2) group (FEP individual or control) and 3) timepoint*group interaction as predictor variables. Cognitive change scores were estimated for each individual as random slopes from their pre-morbid (timepoint 1 and 2) to post-morbid (timepoint 3 and 4) measurements. Akaike Information Criterion (AIC) and Bayesian Information Criterion (BIC) values were used for model comparison at each step as complexity was added to the models.

To specifically assess premorbid to postmorbid changes in the FEP individuals, we rearranged the dummy fixed effect variables to estimate the combined effect of becoming unwell (i.e. timepoints 1 and 2 to timepoints 3 and 4). Because this alternative model is identical in every way except for the construction of the dummy variables, it yields identical predicted values and residuals but allows for interpretation of this combined effect.

The relationship between cognitive change and C4A was explored using multiple linear regression, where age and cognitive change were used to predict levels of C4A. Simple linear regression was used to analyze the relationship between cognitive change and PRS scores.

Cognitive change was also tested as a predictor of several aspects of clinical outcome. Count variables (number of hospital days and number of hospital admissions) were characterized by excess zeroes and were therefore analyzed using hurdle models ^51^. These models allow for simultaneous analysis of the likelihood of having any admissions/hospital days (in the binary component of the model) and the number of admissions/hospital days among those who experienced at least 1 admission/hospital day (truncated count component) ^52^. Distribution of data and model fit (assessed using AIC) was used to guide the choice of statistical model. Logistic regression was used for the binary component of the models and truncated negative binomial models (AIC for truncated Poisson = 9489, AIC for truncated negative binomial = 714) were employed for the count component of the model. In order to explore potential relationships between the degree of cognitive change and long-term likelihood of using psychiatric care for 1), any psychiatric diagnosis, or 2), a main diagnosis of schizophrenia spectrum as well as long-term likelihood of using 3) antipsychotic medications, we used logistic regression models.

The statistical analysis plan was pre-registered on osf.io. All statistical analyses were conducted in R ^53^ version 4.4.0 (2024-04-24). LME models were calculated using the lme4 ^54^ and lmerTest packages ^55^, and hurdle models using the pscl package ^56^. Given the exploratory nature of the study, no corrections for multiple comparisons were made.

## Results

Demographic information is provided for the final sample of 72 FEP individuals and 53 controls in Table 1.

**Table 1.** Demographic comparison of study participants.

|  | Baseline |  |  |  |  | 1.5 year follow up |  |  |  |  |
| --- | --- | --- | --- | --- | --- | --- | --- | --- | --- | --- |
|  | FEP | HC | Chi-2 | t | p | FEP | HC | t | Chi-2 | p |
| N | 72 | 53 |  |  |  | 30 | 23 |  |  |  |
| Age (SD) | 26.6 (6.0) | 26.3 (5.1) |  | -0.34 | 0.737 | 29.0 (6.5) | 27.9 (6.2) | -0.65 |  | 0.516 |
| Sex (female) (%) | 26 (36) | 29 (55) | 3.57 |  | 0.059 | 12 (40) | 13 (57) |  | 0.84 | 0.360 |
| Years of education (SD) | 13.9 (2.8) | 14.7 (2.1) |  | 1.83 | 0.071 | 13.4 (2.5) | 15.9 (2.5) | 3.55 |  | 0.001 |

FEP = Individual with First episode psychosis, HC = healthy control participant, SD = Standard deviation

The two groups were of similar age at inclusion and there were no statistically significant differences in sex composition or years of education at baseline. At follow-up, there was a significant difference in years of education between groups (with FEP participants completing fewer years of education on average) but not for age or sex. Clinical data for individuals with FEP at the baseline assessment and the 1.5-year follow-up can be found in Table 2.

**Table 2.**
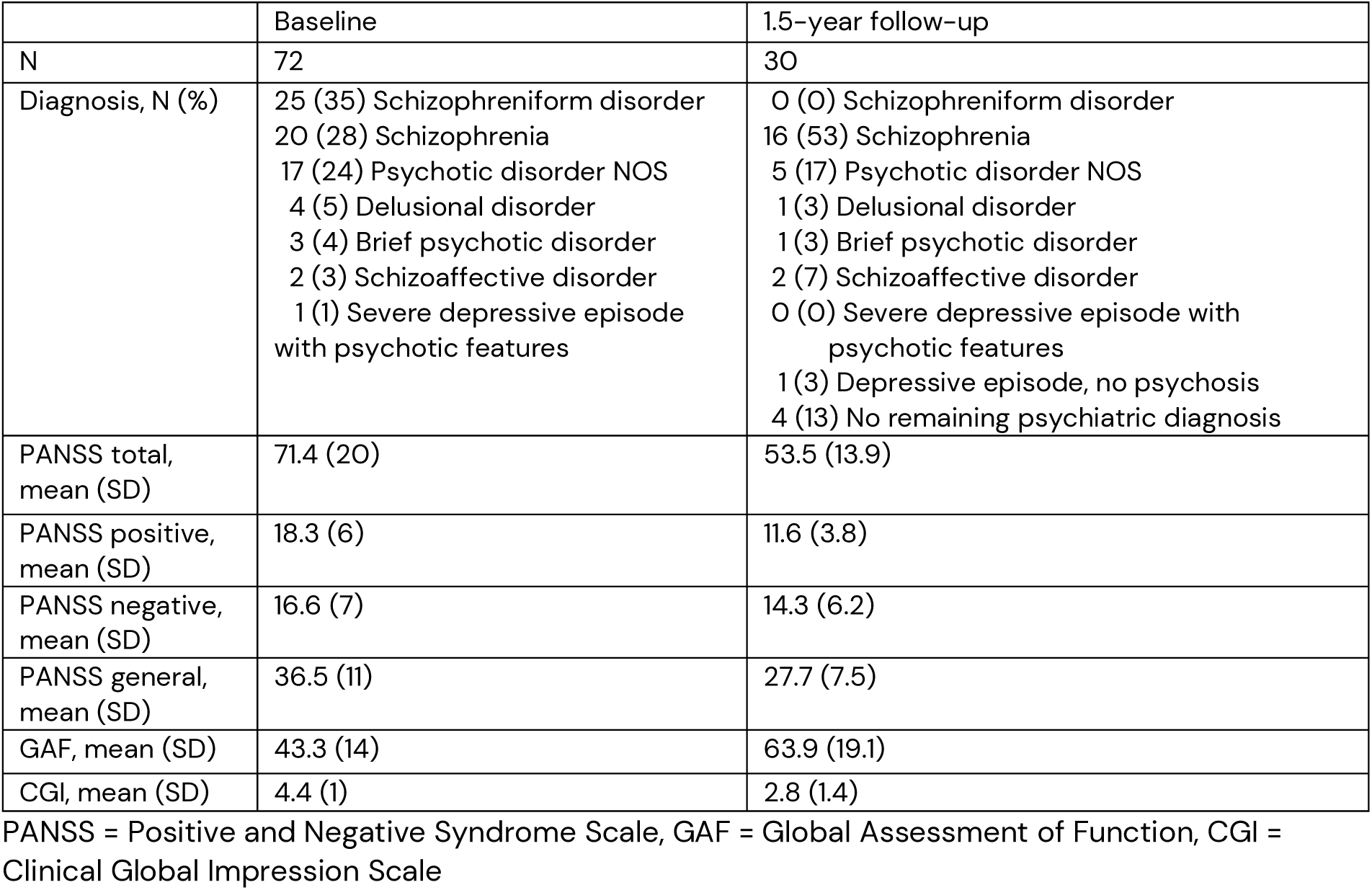
Clinical data for individuals with FEP at baseline and follow-up.

Data on school grades from year 9 were available for 71 patients and all controls, data from year 12 were available for 21 patients and 14 controls. All 72 patients and 53 controls provided data from baseline cognitive testing, and at the 1.5-year follow-up, 30 patients and 23 controls provided cognitive data. Figure 1 shows the available information using boxplots with individual data points superimposed.

**Figure 1.**
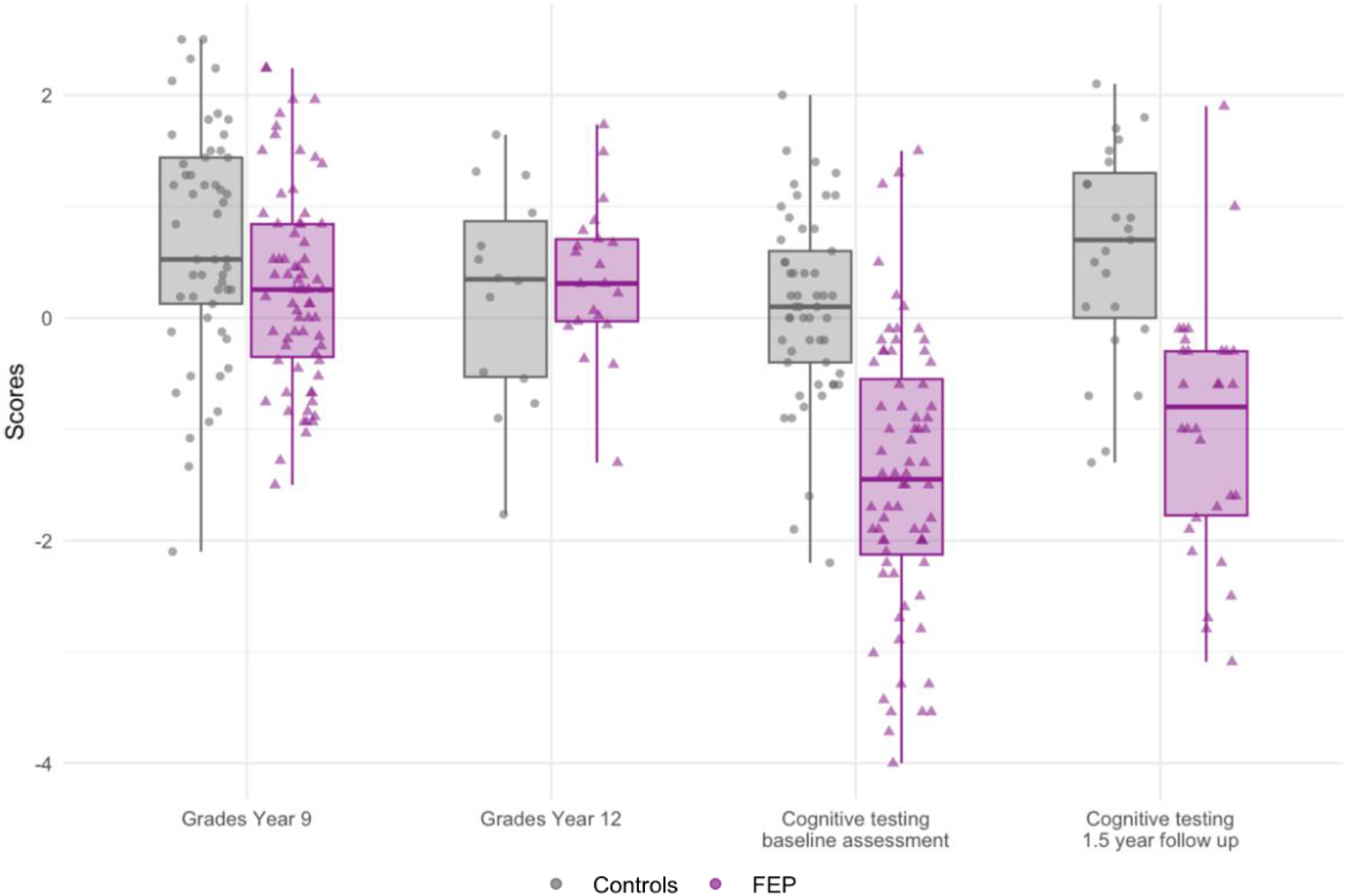
Cognitive data available across timepoints per group

LME models were used to investigate cognitive function over time, defined as either school grades or results from cognitive tests. There was a wide range in years elapsed from completion of grade 9 and inclusion in study (see Supplementary Material, Table 1). Hence, time was coded according to the four assessment phases, 1 through 4. Adding random slopes for patients, to model individual cognitive change from premorbid (school grades) to postmorbid (cognitive testing after illness onset) timepoints, significantly improved model fit (χ2(1) = 35.59, p <.001, AIC/BIC updated model: 856/898). Adding random slopes in a similar fashion for healthy controls further improved model fit (χ2(1) = 12.76, p <.001, AIC/BIC updated model: 845/891) (See Supplementary Material for the final LME model specified). A summary of the final model, containing random intercept, timepoint, group, timepoint*group interaction and random slopes for patients and controls can be found in Table 3.

**Table 3.**
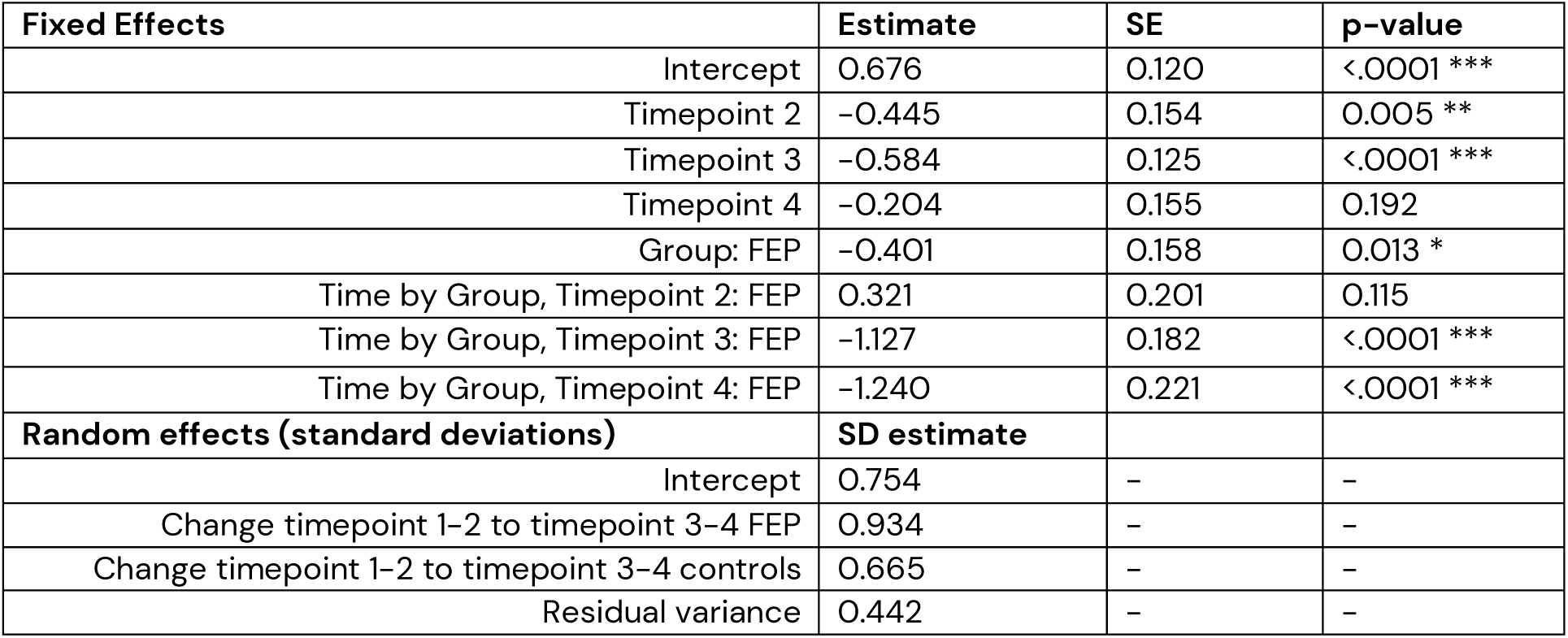
Summary of final LME model of cognitive change across timepoints.

Table 4 presents model-estimated means for each timepoint and contrasts between timepoints for both groups, and this is also depicted graphically in Figure 2A. For individuals with FEP, there was a significant decline between timepoint 2 and timepoint 3, where illness onset was associated with an average loss of 1.12 z-scores: over 1 standard deviation. For controls, there was a significant drop between timepoint 1 and 2, but no significant difference from 2 to 3. Controls significantly improved from baseline cognitive assessment to 1.5-year follow-up, whereas FEP individuals did not.

**Figure 2.**
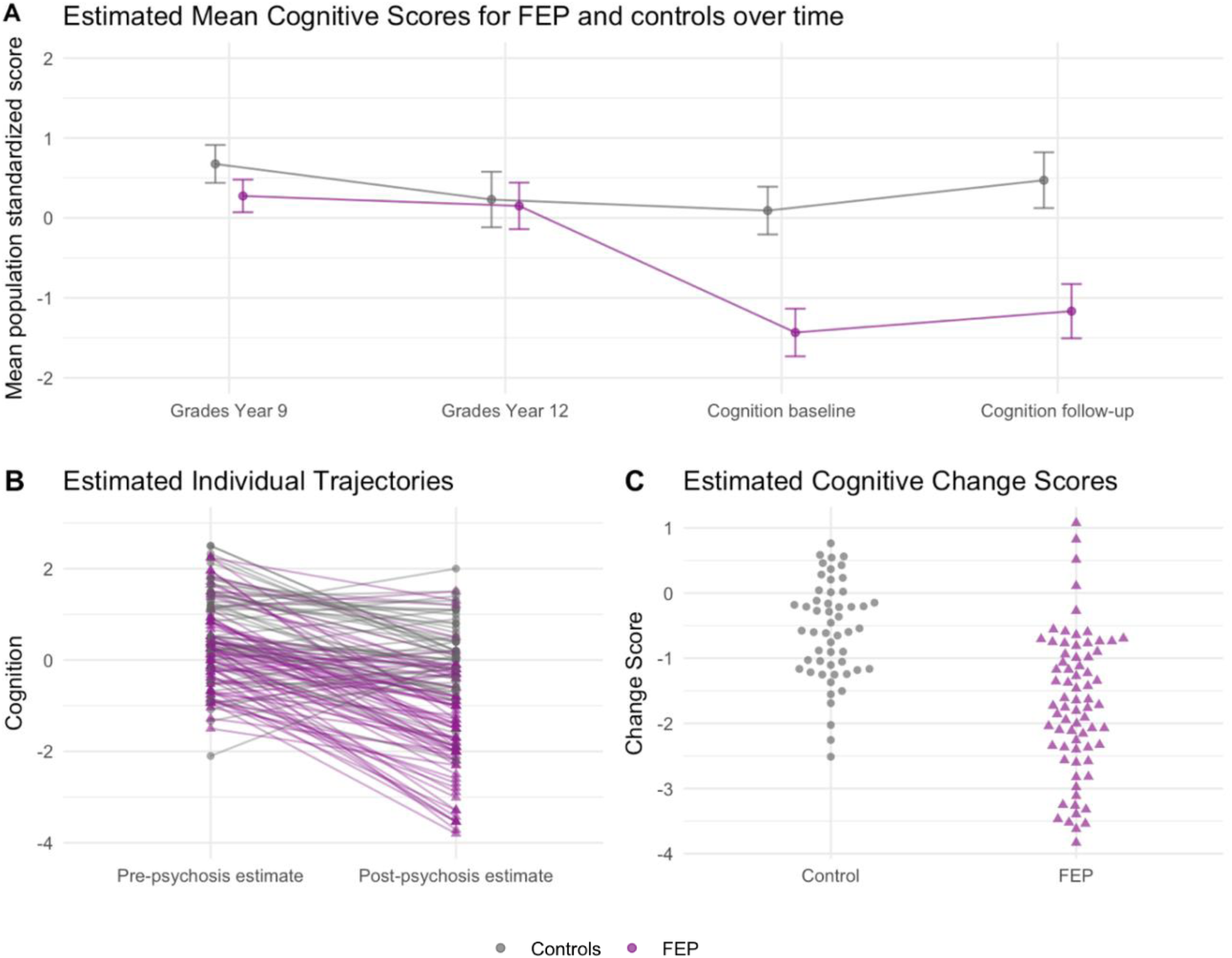
Linear Mixed Effect model estimates

**Table 4.** Model estimated means per timepoint and contrasts between timepoints.

|  | Individuals with FEP |  |  | Controls |  |  |
| --- | --- | --- | --- | --- | --- | --- |
| Mean | Estimate | SE | 95% CI | Estimate | SE | 95 % CI |
| Timepoint 1 | 0.275 | 0.10 | 0.07 – 0.48 | 0.676 | 0.12 | 0.44 – 0.91 |
| Timepoint 2 | 0.151 | 0.15 | -0.14 – 0.44 | 0.230 | 0.18 | -0.12 – 0.58 |
| Timepoint 3 | -1.433 | 0.15 | -1.73 – -1.14 | 0.092 | 0.15 | -0.21 – 0.39 |
| Timepoint 4 | -1.167 | 0.17 | -1.51 – -0.83 | 0.472 | 0.18 | 0.12 – 0.82 |
| Contrasts | Estimate | SE | p | Estimate | SE | p |
| Time2 – Time1 | -0.124 | 0.13 | 0.917 | -0.445 | 0.16 | 0.031 |
| Time3 – Time2 | -1.584 | 0.17 | <.0001 | -0.138 | 0.18 | 0.971 |
| Time4 – Time3 | 0.266 | 0.11 | 0.113 | 0.380 | 0.13 | 0.021 |

In terms of inter-individual variability, the modelled variability estimate for the random slopes was greater in individuals with FEP compared to controls (SD estimate 0.934 vs. 0.665), visualized in Figure 2B-C. The mean effect of becoming unwell in the FEP group was-1.12 (95% CI:-1.48 to-0.77), and the estimated 95% range at the individual level (i.e. after incorporating the random slope variance) was between-2.95 to 0.70.

Panel A: Marginal means plot of estimated mean cognitive scores for individuals with FEP and controls at the four timepoints. Panel B: Estimated individual trajectories in cognitive change from before to after psychosis onset. Panel C: Estimated cognitive change scores in FEP and controls.

### Clinical outcomes

During the study period (from inclusion until 2022-09-30) 79% of patients were admitted to inpatient care due to a psychiatric diagnosis or self-harm at least once. For the patients that were ever hospitalized, the mean number of hospitalizations was 2.8 (median = 2, range = 1-13). The mean number of hospital days (excluding days in mandated involuntary care in outpatient settings) for patients who were ever admitted to inpatient care was 98.2 (median = 40, range 3 – 759). Descriptive statistics and results for the hurdle models are provided in Table 5. There were no associations between the level of cognitive change and the likelihood of ever being hospitalized or number of days spent in hospital for psychiatric reasons.

**Table 5.** Hurdle models of the relationship between cognitive change and hospitalization.

|  | Descriptive | Hurdle model – logistic binary component |  | Descriptive | Hurdle model – truncated negative binomial component |  |
| --- | --- | --- | --- | --- | --- | --- |
|  | No. (%) | OR (95 % CI) | p value | Mean, Median (Range) | IRR (95 % CI) | p value |
| Days in hospital | 57 (79 %) | 0.79 (0.39–1.60) | 0.513 | 98.2, 40, (3 – 759) | 0.97 (0.62–1.52) | 0.905 |
| Number of admissions | 57 (79 %) | 0.79 (0.39–1.60) | 0.513 | 2.8, 2, (1 – 13) | 1.20 (0.69–2.10) | 0.514 |

### Health care usage

Sixty-eight of the 72 eligible patients were followed for approximately 5 years or more (one patient was missing 12 days until 5-year follow up but was also included). For those with sufficient follow-up time, the 12-month prevalence of health care usage in their fifth year after inclusion in the study was summarized and analyzed in relation to cognitive change.

In this long-term follow-up sample, 49 individuals (72%) were still in contact with specialized outpatient psychiatry (at least one visit with a medical doctor).

Thirty-four individuals (50%) had contacts with specialized outpatient psychiatry services where schizophrenia spectrum and other psychotic disorders (ICD-10 codes, F20-F29) were recorded as the main diagnosis. In total, 42 individuals (62%) were still taking antipsychotic medications, based on registered dispensations.

Using logistic regression, we analyzed whether the magnitude of cognitive loss leading up to psychosis could predict subsequent clinical outcomes in the fifth year following inclusion in the study. Cognitive change was not significantly related to whether a person was still in contact with outpatient specialty psychiatry (for any diagnosis); odds ratio (OR) of being a patient at 5 years, per unit decrease in slope-value = 1.27, *p* = 0.471. However, greater cognitive decline was significantly associated with increased likelihood of having an outpatient psychiatry contact for psychotic disorder (OR = 1.89, *p* = 0.046) and with increased likelihood of purchasing antipsychotic medications (OR = 2.08, *p* = 0.027).

#### Biological markers

C4A protein levels were not related to the level of cognitive change in patients (β = 0.017, *p* = 0.384). There was no relationship between genetic risk and cognitive change in patients, for either intelligence PRS (β =-0.132, *p* = 0.372) or schizophrenia PRS (β =-0.005, *p* = 0.978).

#### Sensitivity analysis

When also adjusting for sex, the effect of cognitive change on the likelihood of being a patient in outpatient psychiatry for a psychosis diagnosis was no longer statistically significant (OR = 2.28, p = 0.067), although the effect size did not change substantially. Cognitive change was still a significant predictor of using antipsychotics in the fifth year after inclusion, even when adjusting for sex (OR = 1.96, p = 0.034).

## Discussion

In this study combining registry data on school grades as a proxy of cognitive functioning and results from standardized cognitive tests, we observed a large mean cognitive decline in patients at psychosis onset, with considerable variability in individual trajectories. The magnitude of cognitive change over time predicted long-term clinical outcome, but was not related to number of inpatient days, genetic risk or cerebrospinal fluid levels of C4A.

The average effect associated with illness onset in our sample was large, a loss of over 1.1 population standard deviations, which is in line with previous cross-sectional comparisons with healthy controls ^2,3,25,57^ and longitudinal research in patients ^16^. With regard to inter-individual differences in cognitive decline, the standard deviation among FEP individuals was 0.934, implying a 95% population range of between-2.95 and +0.70. This demonstrates that there are large differences between individuals in the degree of cognitive change in the years between school and psychosis onset, which to our knowledge has not been quantified in this way before.

Cognitive ability did not differ significantly between groups prior to psychosis onset. This contrasts with previous follow-back studies ^21–23^ that have shown differences in school performance before age 16. However, as effect sizes reported in previous studies were small, we may have been unable to detect these differences given our sample size. When examining differences between specific timepoints, there was a general tendency, in both patients and controls (although only statistically significant in controls), for individuals to experience a decline in performance between school years 9 and 12. We believe that this is likely explained by the change in comparison population used to derive the population standardized scores since not all individuals who graduate from year 9 go on to graduate from year 12. Hence, the comparison population only contains those who go on to higher education, while those who do not continue are overrepresented in the lower ranges of the distribution. Furthermore, cognitive performance tended to improve between the baseline assessment at study inclusion and the 1.5 year follow-up in both patients and controls. This improvement might therefore be, at least partly, explained by practice effects ^58^

In terms of clinical outcomes, we were not able to demonstrate any relationship between cognitive change and number of admissions or hospital days, adjusted for time at risk. This finding conflicts with previous research examining associations between cognitive function and hospitalization ^31^. There may be several explanations for this discrepancy. Hospitalizations might be more dependent on a lack of cognitive resources, rather than a relative cognitive loss.

Most FEP individuals in our cohort were recruited from inpatient settings, which is consistent with previous studies in Sweden showing that a majority are admitted to hospital at least once during the first five years of illness ^59^. As such, the first hospitalization may not be as clear of an indicator of poor outcome as any subsequent hospitalizations. For long-term clinical outcome, the level of cognitive decline was not related to likelihood of still being a patient in psychiatry in general; however, it was related to likelihood of being a patient with psychosis diagnosis and likelihood of having dispensed antipsychotic medications. This could indicate that those who experience greater cognitive decline are more likely to need long-term treatment for psychosis.

Cognitive change was not associated with PRS of intelligence or schizophrenia. Effects of these genetic risk markers on clinical outcomes are generally very small ^60–62^, which in combination with the small sample size could explain the lack of correlations. Notably, PRS for cognition and schizophrenia have been shown to differ significantly between cognitive trajectory groups in larger samples ^26^. It could also be speculated that the relationship between cognitive trajectories and PRS depends not only on the relative change from previous levels (which is what we modelled), but also that it is informed by the initial level of cognitive function (which was included in the subgroup analysis implemented in the previous study by Dickinson et al.^26^). Beyond genetic risk, we hypothesized that C4A levels (a marker related to synaptic pruning), would be associated to the degree of cognitive decline. However, we found no evidence of such a relationship in our sample. C4A levels have previously shown to predict development of schizophrenia in our FEP sample ^48^ and higher levels have been associated with worse cognitive performance in larger samples of patients ^63,64^. The lack of a relationship with cognitive change suggests that other mechanisms may be involved in the longitudinal course of cognition than those associated with cognitive impairment when illness has been established.

### Limitations

There are important limitations to acknowledge. While we sought to model more precise trajectories of cognitive change, the large variation in age at psychosis onset made it infeasible to model time in years. Additionally, we were not able to retrieve data for all individuals at all timepoints, with data being particularly sparse at year 12 and 1.5-year follow-up.

The KaSP study was conceptualized to specifically study biomarkers of interest at illness onset. This design allows for unique analysis of such markers very early on; however, as we included drug-naïve and minimally-treated individuals with FEP our cohort may not be fully representative of the broader psychosis patient population. This, along with the fact that substance-use was an exclusion criterion, limits the generalizability of our findings.

There are a few caveats to mention when interpreting clinical outcomes. Firstly, we were not able to control for emigration. This means that individuals missing from medication and care registries might be missing due to emigration, rather than having recovered. Secondly, the medication registry logs dispensations and not prescriptions of drugs. Given the fact that non-adherence to antipsychotics is common ^65^, it is therefore plausible that the registry does not identify all individual in our sample where clinicians have assessed a need for continued treatment. Finally, outpatient care registries only capture visits with medical doctors in specialized care, potentially underestimating the level of outpatient care. However, in specialized psychiatric care for psychosis, there is a mandate to provide yearly follow-ups with a medical doctor to all patients. This was also the reason why we chose to examine the 12-month period prevalence of outpatient contacts.

### Conclusions

Our results demonstrate a large degree of variability between individuals in cognitive change from before to after psychosis onset. The degree of cognitive change was not associated with the selected biological variables but did predict worse clinical outcomes. Given the exploratory nature of the analysis, our results require replication in independent samples.

### Clinical implications

This study demonstrates that individuals with FEP not only exhibit great variability in cognitive impairment at illness onset, but also great variability in relative cognitive decline compared to pre-morbid estimates of cognitive function. This further supports the importance of individual neuropsychological assessments, which should not solely rely on population norms to define impairment but also include estimates or measurements of pre-morbid cognitive function.

## Data Availability

Owing to institutional restrictions, the data cannot be shared openly but can instead be made available upon request on a case-by-case basis as allowed by legislation and ethical permits. Requests for access can be made to the Karolinska Institutets Research Data Office.

## Acknowledgements

We wish to thank the volunteers with FEP and the HC subjects without whom this study had not been possible. We also thank research nurses Joachim Eckerström, Marie Adolfsson, Minna Juntura, Martin Szabo, Henrik Gregemark, Caroline Westerberg-Hake, Lena Lundholm; and the staff at Psykiatri Nordväst, Norra Stockholms Psykiatri, PRIMA Vuxenpsykiatri, Psykiatri Södra, WeMind Psykiatri.

