## Supplementary Material for "Trajectories of Cognitive Function in First-Episode Psychosis: Associations to Clinical Outcomes and Biomarkers"

### Transformation of grades to population standardized scores

In order to compare grades and cognitive tests, these had to be converted to the same scale. Grades from year 9 were available for 124 participants in the KaSP cohort, earliest date 1989 and latest date 2018. Grades from year 12 were available for 35 participants in the KaSP cohort, earliest date 1991 and latest date 2017. The Swedish National Agency for Education also provides summary data on a national level. We utilized the percentile table, where the "grade values" for every 5$^{th}$ percentile for each year is presented. These tables are publicly available from 1998 (grade 9) and 2008 (year 12), and for the years prior we requested similar tables. Summary tables were divided into every 5$^{th}$ percentile, but by writing a function in R (using the approx function, see code available at Github for details) more exact percentile values could be estimated.

Individual participant grades were then converted to percentiles by comparing their results to percentile values for the entire population that graduated in Sweden that year. This was performed for grades from year 9 and year 12, respectively. The percentile scores were then transformed to their corresponding z-scores: this is a one-to-one independent transformation which maps the scores from a bounded uniform to a normal distribution which more accurately summarises the distances between extreme values, and allows the use of parametric statistical models (see flow chart Supplementary Figure).

Of note is that the Swedish national curriculum and grading system has been revised several times, with major revision introduced in 1994 and 2011, as well as a partial revision in 2018 [1]. For the KaSP cohort, this means that individuals who went to school before year 1994 had one grading system (relative grading on a 5 point scale), those in the years 1994 to 2011 had another grading system (goal/knowledge-related grades on a 4 point scale) and those in the years from 2011 onwards had a third type of grading system (goal/knowledge-related grades on a 6 point scale). For this study we used the overall grade-point average. Given that percentile values were available on a national level per year, the same percentile conversion was applicable to all grade-cohorts.

**Supplementary table 1. Time elapsed between grades**

| | N | Mean (SD) | Median | Range |
|---|---|---|---|---|
| Time from year 9 to inclusion FEP | 71 | 11.2 (6.1) | 11 | 2-24 |
| Time from year 9 to inclusion HC | 53 | 11.0 (5.1) | 10 | 3-27 |
| Time from year 12 to inclusion FEP | 21 | 8.6 (6.1) | 11 | -2 - 19 |
| Time from year 12 to inclusion HC | 14 | 7.6 (5.0) | 8 | 1-14 |

Supplementary Figure: Flow Chart

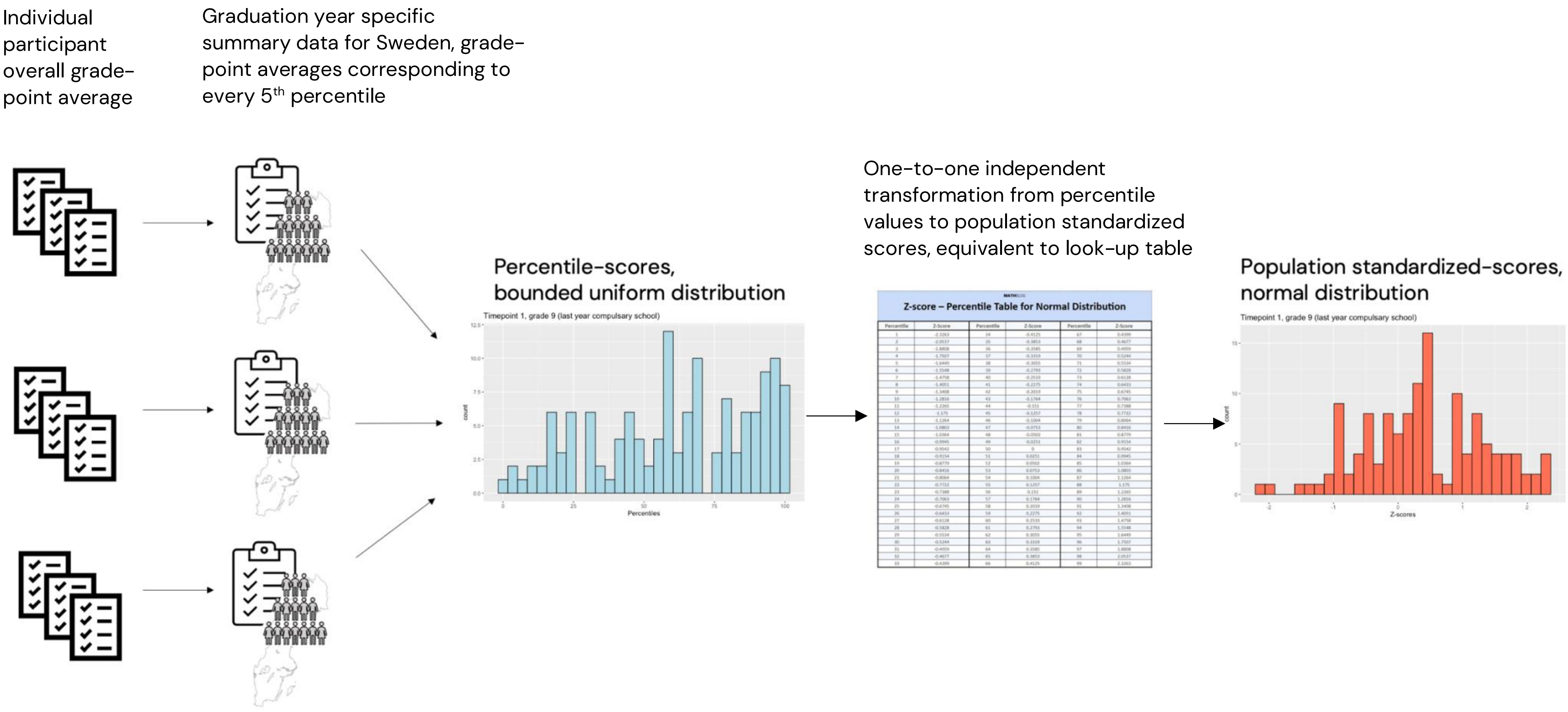

Correlations (Spearman's rho) between school grades and test scores are presented in Supplementary Table 2.

**Supplementary table 2. Correlations between school grades and test scores**

| Group | | Grades year 9 | Grades year 12 | Baseline Assessment | 1.5 year assessment |
|---|---|---|---|---|---|
| FEP | Grades year 9 | - | | | |
| | Grades year 12 | 0.52/ | - | | |
| | Baseline assessment | 0.43 | 0.23 | - | |
| | 1.5 year assessment | 0.45 | -0.01 | 0.89 | - |
| Control | Grades year 9 | - | | | |
| | Grades year 12 | 0.70 | - | | |
| | Baseline assessment | 0.48 | 0.61 | - | |
| | 1.5 year assessment | 0.59 | 0.64 | 0.91 | - |

### Details on Registry data definitions

*Health care usage*

We were interested in health care usage due to psychiatric symptoms and self-harm. In inpatient care (hospitalizations and hospital days) this was primarily defined using the so-called medical activities area, or "Medicinskt Verksamhetsområde" (MVO), codes. These codes identify what kind of health care facility the patient is visiting. All inpatient MVO codes relevant to psychiatry (901, 921, 925, 943, 944, 945) were used. After manually inspecting all the other inpatient spells, two inpatient spells due to intentional self-harm (external cause ICD-codes X7899 and X6409) were added.

Outpatient health care use was also primarily identified using MVO codes, with all codes relevant to psychiatry (581, 901, 906, 921, 923, 925, 928, 929, 931, 943, 944, 945, 948, 951, 954, 955, 956, 957, 991, 993) used to identify specialized outpatient psychiatry visits. In addition to this, visits due to self-harm were identified using external cause ICD-codes (X71 to X83) for intentional self-harm or (X60-X64) for intentional self-poisoning by drugs, medicaments and biological substances. This resulted in 5 more visits for 4 unique patients. Furthermore, visits to other health care facilities, but where the main diagnosis was a psychiatric diagnosis (ICD code starting with F) was also included, resulting in 18 more visits for 10 unique patients.

*Medication dispensation*

Defining anti-psychotic drugs was done by manually inspecting all unique names of drugs dispensed over the follow-up period for the KaSP population available from the Swedish National Prescribed Drug Register (PDR) registry. Initial inspection was conducted by M.L. and inspected by S.C. Mood-stabilizers were not included. Below is the list of the generic names of the dispensed medications that were ultimately defined as anti-psychotics and used in the analysis.

Aripiprazole
Clozapine
Flupentixol
Haloperidol
Klorprotixen
Kvetiapin
Latuda
Levomepromazin
Olanzapin
Paliperidon
Perfenazin
Reagila
Risperidon
Ziprasidon
Zuklopentixol

**Linear Mixed Effect model details**

Specification of Linear Mixed Effects model used in final analysis (lmer syntax):

(Cognitive score ~ 1 + Timepoint*Group +
(1 | ID) + ( 0 + Change for FEP | ID) + (0 + Change for controls | ID)

DATA AVAILABILITY
Owing to institutional restrictions, the data cannot be shared openly but can instead be made available upon request on a case-by-case basis as allowed by the legislation and ethical permits. Requests for access can be made to the Karolinska Institutet's Research Data Office at.

CODE AVAILABILITY
The code for reproducing the analyses and figures in this article will be made available at https://github.com/MariaLeeR/Cognitive_trajectoriesFEP.
